# Detection of Endothelial Plaque in Microbial Keratitis Using Anterior Segment Optical Coherence Tomography

**DOI:** 10.64898/2026.02.03.26345494

**Authors:** Folahan Ibukun, Kamini Reddy, Subeesh Kuyyadiyil, Elesh Jain, Gautam Parmar, Nakul S. Shekhawat

## Abstract

**Purpose:** To evaluate the intra- and inter-grader concordance of anterior segment optical coherence tomography (ASOCT) grading for detection of endothelial plaque in microbial keratitis, and to compare endothelial plaque detection via ASOCT grading versus in-person slit lamp examination.

**Methods:** Diagnostic concordance study of 150 consecutive patients with microbiologically confirmed bacterial or fungal keratitis at a high-volume tertiary eye hospital in India. Two masked ophthalmologist graders independently evaluated ASOCT images for presence of two morphologically distinct endothelial plaque subtypes noted during image review (round and flat plaques). We assessed intra-grader and inter-grader concordance for each endothelial plaque morphology and for presence of either morphology. Diagnostic agreement between ASOCT and in-person slit lamp examination was evaluated using percent agreement, Cohen’s kappa, sensitivity, and specificity. Univariable and multivariable logistic regression was used to assess odds of disagreement between ASOCT and slit lamp examination for endothelial plaque detection.

**Results:** ASOCT detection showed near perfect inter-grader agreement for round endothelial plaques (kappa 0.88, 94.7% agreement), flat endothelial plaques (kappa 0.84, 92.0% agreement), and either plaque (kappa 0.88, 94.0% agreement). Intra-grader agreement was substantial to near perfect for both graders across all plaque types (kappa 0.70-0.86). Ophthalmologist slit lamp examination identified endothelial plaque in 6.0% eyes, while ASOCT detected round plaques in 32.7%, flat plaques in 43.3%, and either plaque in 55.3% of eyes. Using ASOCT as reference, slit lamp examination demonstrated sensitivity of 16.3% for round plaques, 6.2% for flat plaques, and 9.6% for either plaque, with specificity exceeding 94% for all. Poor visual acuity (logMAR ≥1.0) was associated with increased disagreement for round plaques (adjusted OR 5.04), flat plaques (adjusted OR 3.63), and either plaque (adjusted OR 3.98). Bacterial infection was associated with increased disagreement for any endothelial plaque (adjusted OR 4.56).

**Conclusion:** Slit lamp examination substantially under-detects endothelial plaque compared to ASOCT, while ASOCT enables reproducible detection with excellent intra- and inter-grader agreement. These findings support incorporation of ASOCT imaging into microbial keratitis evaluation protocols. Differences in round and flat endothelial plaque morphologies warrant further investigation.

## INTRODUCTION

Microbial keratitis is a leading cause of corneal blindness worldwide, with an estimated 1.5 to 2 million cases occurring annually.^1,2^ The global burden disproportionately affects developing countries, where occupational injuries from agricultural work and limited access to eye care contribute to both higher infection incidence and poorer outcomes.^3^ The etiology of microbial keratitis varies by geographic region, with bacterial infections predominating in developed countries and fungal keratitis accounting for 30-60% of cases in tropical and agricultural regions.^1,4,5^ Regardless of causative organism, severe microbial keratitis can progress rapidly to corneal perforation, endophthalmitis, and permanent vision loss.

Endothelial plaque, a retrocorneal inflammatory deposit adherent to the posterior corneal surface, represents an important clinical finding in microbial keratitis. Endothelial plaques are composed of inflammatory cells, fibrin, and in fungal infections may contain viable organisms that have penetrated through Descemet membrane into the anterior chamber.^6,7^ The presence of endothelial plaque has been associated with more severe disease presentation, larger infiltrate size, worse visual outcomes, and need for procedural intervention.^6–10^ In fungal keratitis specifically, endothelial plaques may indicate anterior chamber invasion of the infectious organism^6,8^ and have been proposed as a predictor of treatment failure and need for surgical intervention.

Despite their clinical significance, endothelial plaques can be challenging to detect on routine slit lamp examination. Dense stromal infiltrates, corneal edema and haze, and hypopyon may obscure visualization of the posterior cornea and endothelial surface. This limitation is particularly problematic in severe infections where accurate assessment of disease extent is most critical for guiding management decisions. Anterior segment optical coherence tomography (ASOCT) provides high-resolution cross-sectional imaging of corneal anatomy and has emerged as a valuable adjunct for evaluating microbial keratitis.^7,10^ ASOCT can visualize corneal layers, quantify infiltrate depth and stromal thinning, and potentially detect endothelial plaques that may not be apparent on clinical examination.

The reliability of ASOCT for detecting endothelial plaques in microbial keratitis has not been systematically evaluated. Furthermore, endothelial plaques may present with different morphologies and response to treatment that could have distinct pathophysiologic mechanisms and clinical implications. Understanding the reproducibility and diagnostic performance of ASOCT for detecting different endothelial plaque morphologies is essential before incorporating ASOCT into clinical protocols or research studies for microbial keratitis. The purpose of this study was to evaluate the intra-grader reproducibility, inter-grader concordance, and diagnostic agreement of ASOCT with ophthalmologist slit lamp examination for detection of endothelial plaques in eyes with microbiologically confirmed microbial keratitis.

## METHODS

### Study Design and Population

We conducted a prospective diagnostic concordance study among patients with microbiologically confirmed keratitis treated at SNC Chitrakoot Eye Hospital, Chitrakoot, Madhya Pradesh, India, between May 2024 and January 2025. Inclusion criteria were clinical diagnosis of microbial keratitis, confirmation of bacterial or fungal keratitis by corneal scraping and smear microscopy, and availability of ASOCT imaging upon clinical presentation. Patients were classified as having bacterial, fungal, or polymicrobial keratitis based on microbiological results. The study adhered to the tenets of the Declaration of Helsinki and was approved by the Johns Hopkins University School of Medicine Institutional Review Board and the SNC Chitrakoot research ethics committee.

### Clinical Examination and Imaging

All patients presenting for their initial visit to the hospital underwent comprehensive ophthalmic examination including visual acuity measurement, ophthalmologist-performed slit lamp biomicroscopy with standardized notation of examination findings, slit lamp photography with diffuse and blue light illumination, and ASOCT imaging using the Heidelberg Anterion platform’s Metrics application (Heidelberg Engineering, Heidelberg, Germany). The Anterion Metrics app obtains six radially oriented scans at 0, 30, 60, 90, 120, and 150 degrees, enabling limbus-to-limbus assessment of corneal and anterior segment anatomy at each of these meridians. The examining ophthalmologist recorded the presence or absence of endothelial plaque on the clinical examination form. For each eye, we created a four-panel composite image comprising the ASOCT scan, grayscale reference photograph showing the angle at which the ASOCT scan cut was taken, slit lamp photograph with diffuse illumination, and slit lamp photograph with cobalt blue light taken 15 seconds after instillation of sodium fluorescein dye. Imaging was performed in a separate room from the examination area to prevent biasing ophthalmologists’ slit lamp assessment, and in-person ophthalmologist examiners did not review imaging results before slit lamp examination.

### Remote Grading Protocol

Two trained ophthalmologist image graders (NS, FI) masked to in-person slit lamp examination findings, microbiology testing results, and to the other image grader’s assessment independently evaluated each eye’s six composite images for endothelial plaque. Graders classified lesions into two morphological subtypes: (1) Round plaques, defined as discrete elevated lesions with rounded curvature, indistinct edges, wider diameter at the center of the lesion than at the plane of endothelial attachment, and deeper anterior chamber invasion; and (2) Flat plaques, defined as linear hyperreflective lesions oriented in the same plane as Descemet membrane with sharper margins, minimal anterior chamber protrusion, and broader attachment to the corneal endothelium. Each morphology was graded as present or absent after reviewing all six images per eye. Disagreements between graders were resolved through discussion until a consensus grade was achieved for each eye. To assess intra-grader reproducibility, a random subsample of 45 eyes (30%) underwent repeat grading by each grader after a minimum two-week washout period.

### Statistical Analysis

Separate analyses were performed for round endothelial plaque, flat endothelial plaque, and any endothelial plaque. We calculated intra-grader reproducibility using Cohen’s kappa statistic as well as percent agreement across repeated assessments for the binary classification of presence versus absence of each endothelial plaque category. Inter-grader agreement between the two graders was measured using percent agreement and Cohen’s kappa. Kappa statistic was graded as poor (<0.20), fair (0.21–0.40), moderate (0.41–0.60), substantial (0.61–0.80), or near perfect agreement (0.81–1.00).^11^

Using the two graders’ consensus grade from ASOCT image grading as the reference standard, we estimated diagnostic validity compared to in-person ophthalmologist slit lamp examination. We calculated sensitivity, specificity, positive predictive value (PPV), negative predictive value (NPV), percent agreement, and kappa statistic for each of the three endothelial plaque categories. Of note, slit lamp examination recorded only “endothelial plaque” without distinguishing between round and flat morphologies.

To determine whether endothelial plaque detection in ASOCT versus slit lamp examination varied across different types of infections, subgroup analyses evaluated concordance between ASOCT and slit lamp examination by visual acuity (logarithm of the minimum angle of resolution [logMAR] <1.0 versus ≥1.0), infection type (bacterial, fungal, or polymicrobial), infiltrate/scar diameter (0 to <2mm, 2 to <6mm, ≥6mm), infiltrate depth within the stroma (anterior ⅓, middle ⅓, and/or posterior ⅓ stroma), presence or absence of stromal thinning on slit lamp examination, and infiltrate location within 2mm of the corneoscleral limbus. To identify factors associated with disagreement between remote ASOCT grading and in-person slit lamp examination, we performed univariable and multivariable logistic regression analyses separately for each outcome (round plaques, flat plaques, and either morphology). Multivariable models adjusted for visual acuity, infection type, infiltrate diameter, infiltrate depth, stromal thinning, and infiltrate proximity to the limbus. All statistical analyses were conducted using Stata SE version 18.5. Confidence intervals for diagnostic statistics were derived using bootstrap resampling with 1000 iterations, and percentile-based confidence intervals are presented.

## RESULTS

### Study Population

A total of 150 eyes of 150 patients with microbiologically confirmed bacterial or fungal keratitis met eligibility criteria and were included in the study (**Table 1**). The median patient age was 50.0 years (interquartile range [IQR] 41.0-59.0), and the majority were male (92/150, 61.3%). Fungal keratitis was the most common etiology (80/150, 53.3%) in this heavily agricultural population, followed by bacterial (54/150, 36.0%) and polymicrobial keratitis with both bacterial and fungal infection (16/150, 10.7%). The median logMAR visual acuity was 1.39 (IQR 0.6-2.0), with the majority presenting with visual acuity 20/200 or worse (logMAR ≥1.0 in 120 of 150 eyes, 80.0%) and 52 eyes (34.7%) presenting with light perception only or no light perception. Median duration from symptom onset to presentation to the tertiary care center was 15.0 days (IQR 7.0-30.0). The stromal infiltrate diameter was 2 to <6 mm in 104 eyes (69.3%) and >6 mm in 24 eyes (16.0%). Stromal thinning was present in 46 eyes (30.7%), and infiltrates were located within 2 mm of the limbus in 13 eyes (8.7%).

**Table 1.** Demographic and clinical characteristics of study population.

| **Characteristics** | **Value** |
| --- | --- |
| **Age, median (IQR)** | 50.0 (41.0–59.0) |
| **Sex** |  |
| Female, N (%) | 58 (38.7) |
| Male, N (%) | 92 (61.3) |
| **Eye laterality** |  |
| Right eye, N (%) | 96 (64.0) |
| Left eye, N (%) | 54 (36.0) |
| **LogMAR visual acuity, median (IQR)** | 1.39 (0.6–2.0) |
| LogMAR <1, N (%) | 30 (20.0) |
| LogMAR ≥1, N (%)<sup>a</sup> | 120 (80.0) |
| **Snellen visual acuity, N (%)** |  |
| 6/6 to ≤ 6/18 | 17 (11.3) |
| 6/24 to ≤ 6/36 | 13 (8.7) |
| 1/60 to ≤6/60 | 32 (21.3) |
| Finger counts or Hand movements | 36 (24.0) |
| Light perception or No light perception | 52 (34.7) |
| **Lens Status, N (%)** |  |
| Mature cataract | 1 (0.7) |
| Immature cataract | 31 (20.7) |
| Early lens changes | 31 (20.7) |
| Clear crystalline lens | 57 (38.0) |
| Aphakia | 0 (0.0) |
| Pseudophakia | 8 (5.3) |
| Unable to determine lens status | 22 (14.7) |
| **Days from symptom onset to presentation, median (IQR)** | 15.0 (7.0–30.0) |
| **Infection Type** |  |
| Fungal Only | 80 (53.3) |
| Bacterial Only | 54 (36.0) |
| Polymicrobial | 16 (10.7) |
| **Infiltrate Diameter** |  |
| Not applicable | 1 (0.7) |
| 0 to <2mm | 21 (14.0) |
| 2 to <6mm | 104 (69.3) |
| ≥6mm | 24 (16.0) |
| **Infiltrate Depth** |  |
| Anterior 1/3 Stromal Involvement | 133 (88.7) |
| Middle 1/3 Stromal Involvement | 97 (64.7) |
| Posterior 1/3 Stromal Involvement | 39 (26.0) |
| **Stromal Thinning Present** |  |
| Yes | 46 (30.7) |
| No | 104 (69.3) |
| **Infiltrate Within 2mm of Limbus** |  |
| Yes | 13 (8.7) |
| No | 137 (91.3) |

<sup>a</sup> Includes 51 eyes with light perception and 1 eye with no light perception
Abbreviations : IQR = Interquartile range; logMAR = logarithm of minimum angle of resolution ; N = number

### Intra-Grader and Inter-Grader Concordance of ASOCT Grading

Inter-grader agreement for ASOCT grading was near perfect across all endothelial plaque categories (**Table 2**). For round plaques, agreement was 94.7% (95% CI: 90.7-98.0%) with kappa of 0.88 (95% CI: 0.79-0.95). For flat plaques, agreement was 92.0% (95% CI: 87.3-96.0%) with kappa of 0.84 (95% CI: 0.73-0.92). For either endothelial plaque morphology, agreement was 94.0% (95% CI: 90.0-97.3%) with kappa of 0.88 (95% CI: 0.80-0.95).

**Table 2.**
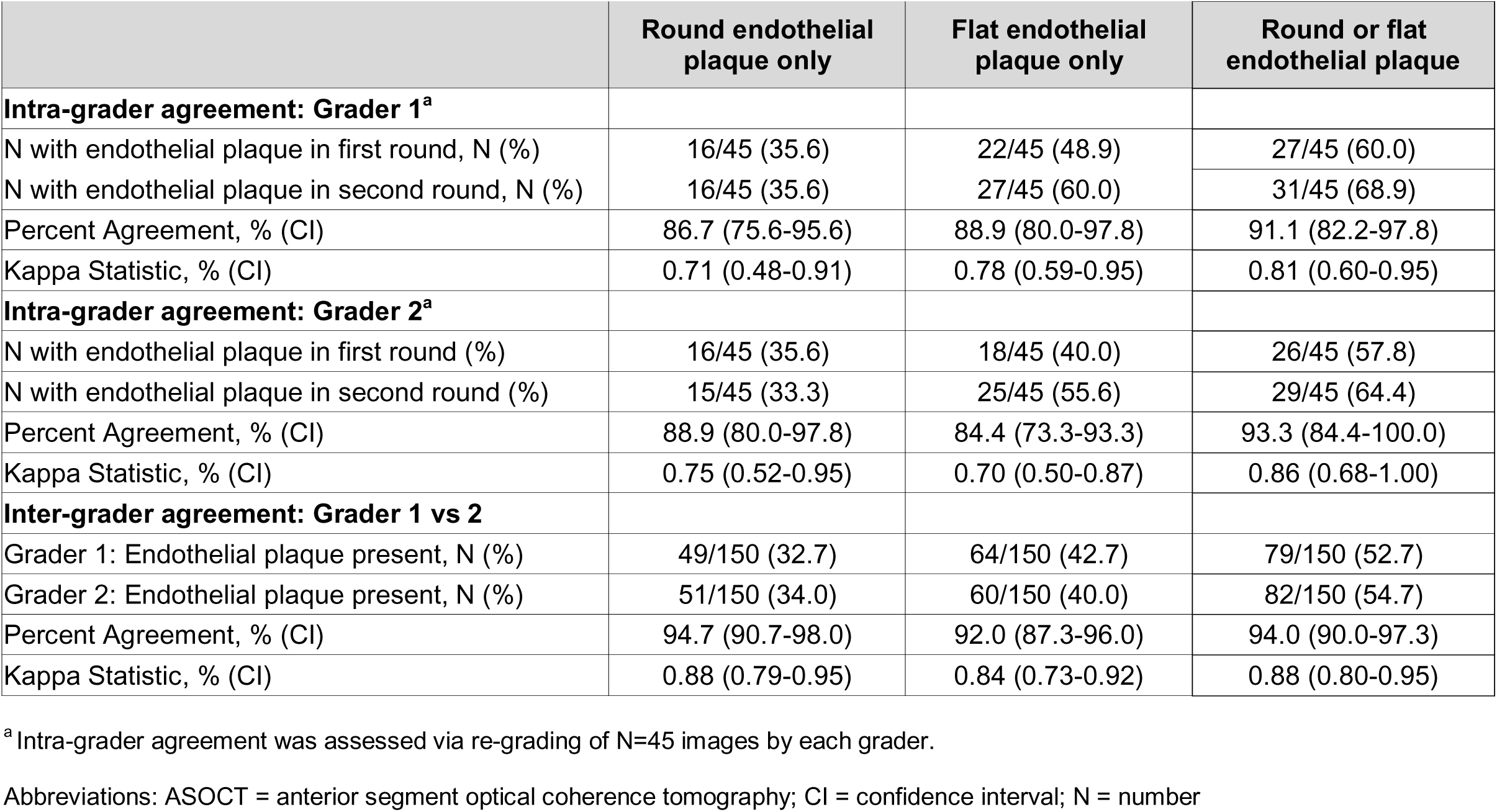
Intra-grader and Inter-grader Agreement of ASOCT for Detection of Endothelial Plaque.

|  | Round endothelial plaque only | Flat endothelial plaque only | Round or flat endothelial plaque |
| --- | --- | --- | --- |
| **Intra-grader agreement: Grader 1<sup>a</sup>** |  |  |  |
| N with endothelial plaque in first round, N (%) | 16/45 (35.6) | 22/45 (48.9) | 27/45 (60.0) |
| N with endothelial plaque in second round, N (%) | 16/45 (35.6) | 27/45 (60.0) | 31/45 (68.9) |
| Percent Agreement, % (CI) | 86.7 (75.6-95.6) | 88.9 (80.0-97.8) | 91.1 (82.2-97.8) |
| Kappa Statistic, % (CI) | 0.71 (0.48-0.91) | 0.78 (0.59-0.95) | 0.81 (0.60-0.95) |
| **Intra-grader agreement: Grader 2<sup>a</sup>** |  |  |  |
| N with endothelial plaque in first round (%) | 16/45 (35.6) | 18/45 (40.0) | 26/45 (57.8) |
| N with endothelial plaque in second round (%) | 15/45 (33.3) | 25/45 (55.6) | 29/45 (64.4) |
| Percent Agreement, % (CI) | 88.9 (80.0-97.8) | 84.4 (73.3-93.3) | 93.3 (84.4-100.0) |
| Kappa Statistic, % (CI) | 0.75 (0.52-0.95) | 0.70 (0.50-0.87) | 0.86 (0.68-1.00) |
| **Inter-grader agreement: Grader 1 vs 2** |  |  |  |
| Grader 1: Endothelial plaque present, N (%) | 49/150 (32.7) | 64/150 (42.7) | 79/150 (52.7) |
| Grader 2: Endothelial plaque present, N (%) | 51/150 (34.0) | 60/150 (40.0) | 82/150 (54.7) |
| Percent Agreement, % (CI) | 94.7 (90.7-98.0) | 92.0 (87.3-96.0) | 94.0 (90.0-97.3) |
| Kappa Statistic, % (CI) | 0.88 (0.79-0.95) | 0.84 (0.73-0.92) | 0.88 (0.80-0.95) |

<sup>a</sup> Intra-grader agreement was assessed via re-grading of N=45 images by each grader.
Abbreviations: ASOCT = anterior segment optical coherence tomography; CI = confidence interval; N = number

Intra-grader reproducibility was substantial to near perfect for both graders across all lesion types. For round plaques, Grader 1 (NS) demonstrated 86.7% agreement (kappa 0.71) and Grader 2 (FI) demonstrated 88.9% agreement (kappa 0.75). For flat plaques, Grader 1 showed 88.9% agreement (kappa 0.78) and Grader 2 showed 84.4% agreement (kappa 0.70). For either morphology, Grader 1 achieved 91.1% agreement (kappa 0.81) and Grader 2 achieved 93.3% agreement (kappa 0.86). Notably, the composite outcome (either round or flat plaque morphology) showed the highest intra-grader reproducibility for both graders.

### ASOCT Versus Slit Lamp Examination

Detection for each of the two graders was similar. Grader 1 identified round plaques in 32.7% and flat plaques in 42.7% of eyes, while Grader 2 identified round plaques in 34.0% and flat plaques in 40.0% of eyes. On consensus remote grading, round endothelial plaques were identified in 49 of 150 eyes (32.7%), flat endothelial plaques in 65 eyes (43.3%), and any endothelial plaque in 83 eyes (55.3%).

By contrast, in-person slit lamp examination detected any form of endothelial plaque in only 9/150 eyes (6.0%; **Table 3**). Percent agreement between ASOCT and slit lamp examination was 72.0% for round plaques, 56.0% for flat plaques, and 49.3% for either morphology. Kappa values showed poor agreement of 0.19 (95% CI: 0.07-0.32) for round plaques, 0.00 (95% CI: -0.09 to 0.09) for flat plaques, and 0.07 (95% CI: 0.01-0.15) for either morphology. The near-zero kappa for flat plaques indicated essentially no agreement beyond chance alone, reflecting the particularly limited detection of flat plaques on slit lamp examination. Using ASOCT as the reference standard, slit lamp examination demonstrated low sensitivity, with 16.3% sensitivity (95% CI: 6.3-26.9%) for round plaques, 6.2% (95% CI: 1.4-12.5%) for flat plaques, and 9.6% (95% CI: 3.6-16.4%) for either morphology. Slit lamp examination showed high specificity, with 99.0% specificity (95% CI: 96.7-100%) for round plaques, 94.1% (95% CI: 88.0-98.7%) for flat plaques, and 98.5% (95% CI: 94.7-100%) for either morphology.

**Table 3.**
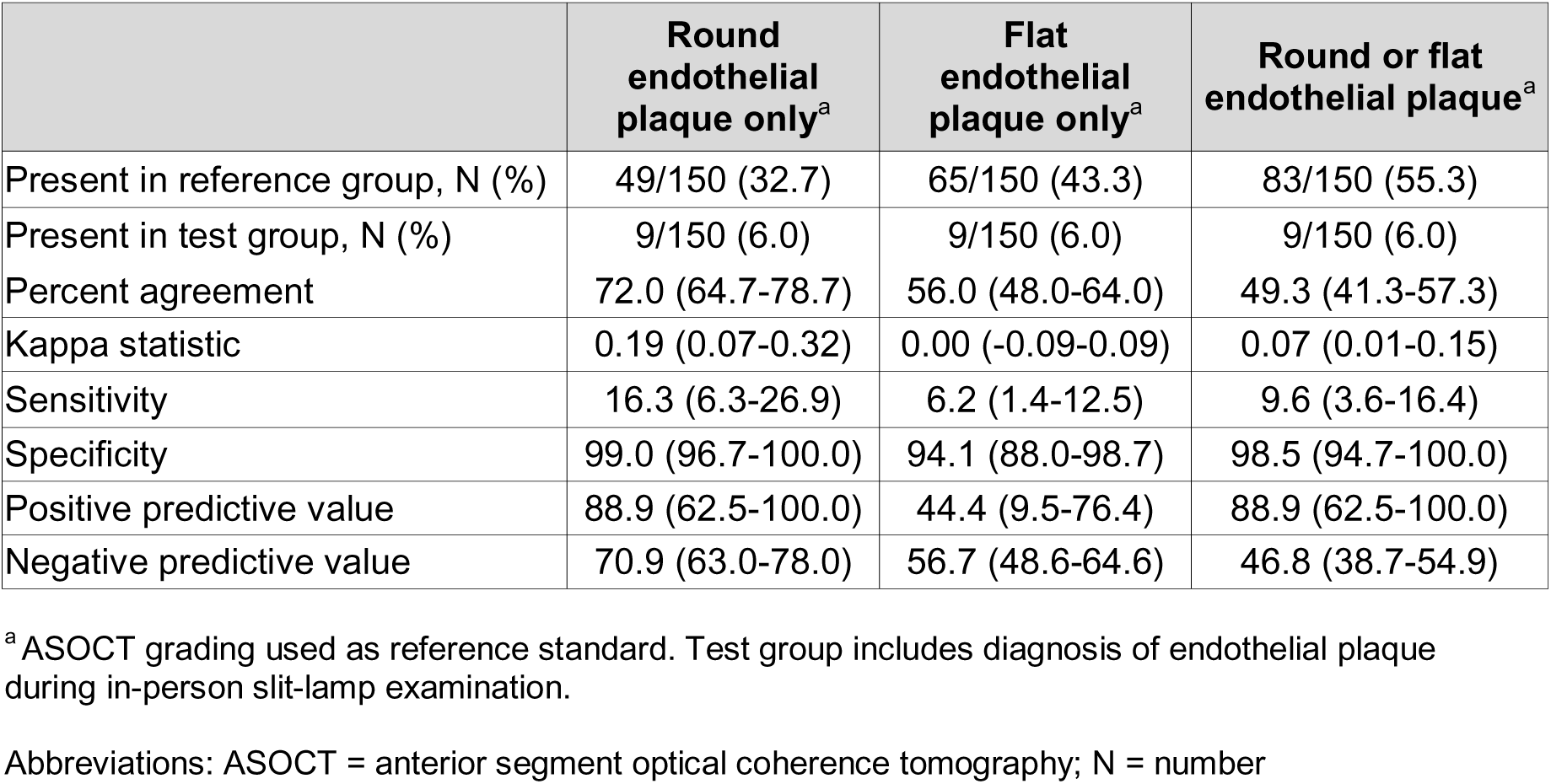
Concordance of ASOCT and Ophthalmologist Slit Lamp Examination for Detection of Endothelial Plaque.

### Factors Associated with Disagreement Between ASOCT and Slit Lamp Examination

When evaluating detection of round endothelial plaque, univariable logistic regression (**Table 4**) showed that worse visual acuity (logMAR ≥1.0) was significantly associated with disagreement between ASOCT and slit lamp examination (OR 7.0, 95% CI: 1.58-31.02). Infiltrate characteristics associated with disagreement included larger diameter (2 to <6mm vs <2mm: OR 9.30, 95% CI: 1.19-72.74), posterior ^1^/_3_ stromal involvement (OR 3.62, 95% CI: 1.67-7.93), middle ^1^/_3_ stromal involvement (OR 3.04, 95% CI: 1.28-7.19), and presence of stromal thinning on slit lamp examination (OR 2.87, 95% CI: 1.35-6.09). On multivariable analysis, poor visual acuity remained significantly associated with disagreement (adjusted OR 5.04, 95% CI: 1.04-24.47) after adjusting for the aforementioned covariates.

**Table 4.**
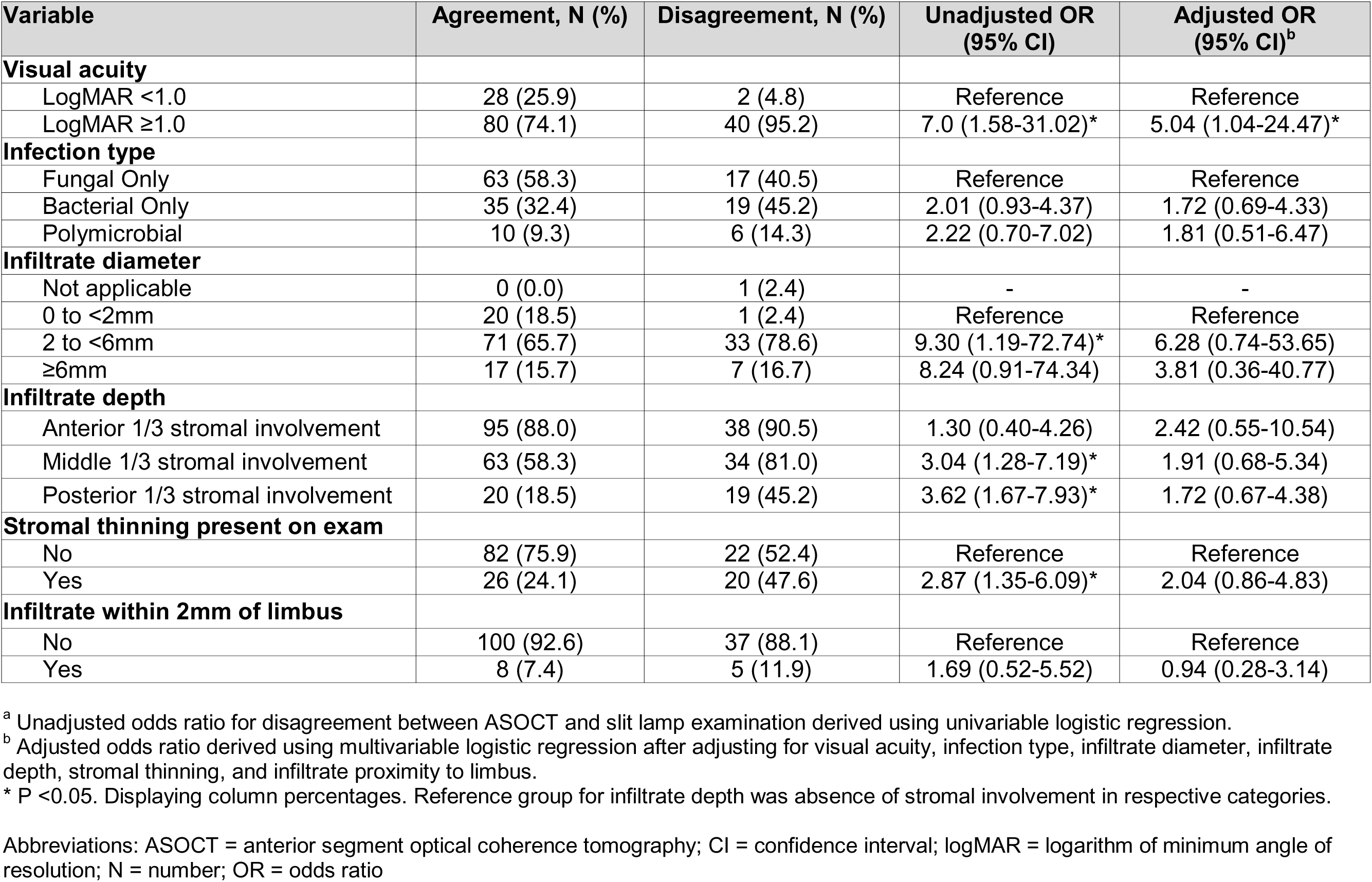
Odds of Disagreement for Round Endothelial Plaque Detection Between ASOCT and Slit Lamp Examination.

When evaluating detection of flat endothelial plaque on ASOCT versus slit lamp examination (**Table 5**), worse visual acuity was significantly associated with disagreement (OR 4.00, 95% CI: 1.52-10.52). Bacterial infection showed increased odds of disagreement compared to fungal infection (OR 2.25, 95% CI: 1.11-4.55), and presence of stromal thinning was associated with disagreement (OR 2.37, 95% CI: 1.16-4.82). On multivariable analysis, poor visual acuity remained significantly associated with disagreement (adjusted OR 3.63, 95% CI: 1.29-10.21).

**Table 5.** Odds of Disagreement for Flat Endothelial Plaque Detection Between ASOCT and Slit Lamp Examination.

| Variable | Agreement, N (%) | Disagreement, N (%) | Unadjusted OR (95% CI) | Adjusted OR (95% CI)<sup>b</sup> |
| --- | --- | --- | --- | --- |
| **Visual acuity** |  |  |  |  |
| LogMAR <1.0 | 24 (28.6) | 6 (9.1) | Reference | Reference |
| LogMAR ≥1.0 | 60 (71.4) | 60 (90.9) | 4.00 (1.52-10.52)* | 3.63 (1.29-10.21)* |
| **Infection type** |  |  |  |  |
| Fungal Only | 50 (59.5) | 30 (45.5) | Reference | Reference |
| Bacterial Only | 23 (27.4) | 31 (47.0) | 2.25 (1.11-4.55)* | 1.94 (0.90-4.18) |
| Polymicrobial | 11 (13.1) | 5 (7.6) | 0.76 (0.24-2.40) | 0.75 (0.22-2.54) |
| **Infiltrate diameter** |  |  |  |  |
| Not applicable | 1 (1.2) | 0 (0.0) | - |  |
| 0 to <2mm | 13 (15.5) | 8 (12.1) | Reference | Reference |
| 2 to <6mm | 57 (67.9) | 47 (71.2) | 1.34 (0.51-3.52) | 1.36 (0.46-4.01) |
| ≥6mm | 13 (15.5) | 11 (16.7) | 1.38 (0.42-4.55) | 0.97 (0.25-3.80) |
| **Infiltrate depth** |  |  |  |  |
| Anterior 1/3 stromal involvement | 75 (89.3) | 58 (87.9) | 0.87 (0.32-2.40) | 1.17 (0.37-3.71) |
| Middle 1/3 stromal involvement | 51 (60.7) | 46 (69.7) | 1.49 (0.75-2.95) | 1.25 (0.54-2.91) |
| Posterior 1/3 stromal involvement | 19 (22.6) | 20 (30.3) | 1.49 (0.71-3.10) | 0.75 (0.30-1.89) |
| **Stromal thinning present on exam** |  |  |  |  |
| No | 65 (77.4) | 39 (59.1) | Reference | Reference |
| Yes | 19 (22.6) | 27 (40.9) | 2.37 (1.16-4.82)* | 1.90 (0.84-4.29) |
| **Infiltrate within 2mm of limbus** |  |  |  |  |
| No | 77 (91.7) | 60 (90.9) | Reference | Reference |
| Yes | 7 (8.3) | 6 (9.1) | 1.10 (0.35-3.46) | 0.79 (0.21-2.99) |

<sup>a</sup> Unadjusted odds ratio for disagreement between ASOCT and slit lamp examination derived using univariable logistic regression.
<sup>b</sup> Adjusted odds ratio derived using multivariable logistic regression after adjusting for visual acuity, infection type, infiltrate diameter, infiltrate depth, stromal thinning, and infiltrate proximity to limbus.
\* P <0.05. Displaying column percentages. Reference group for infiltrate depth was absence of stromal involvement in respective categories.
Abbreviations: ASOCT = anterior segment optical coherence tomography; CI = confidence interval; logMAR = logarithm of minimum angle of resolution; N = number; OR = odds ratio

When evaluating detection of either type of endothelial plaque (**Table 6**), poor visual acuity was significantly associated with disagreement between ASOCT and slit lamp examination on univariable analysis (OR 4.44, 95% CI: 1.77-11.19). Bacterial infection showed the strongest association with disagreement (OR 4.57, 95% CI: 2.15-9.70), and presence of stromal thinning was associated with disagreement (OR 3.60, 95% CI: 1.69-7.65). On multivariable analysis, both poor visual acuity (adjusted OR 3.98, 95% CI: 1.32-11.98) and bacterial infection (adjusted OR 4.56, 95% CI: 1.97-10.57) remained significantly associated with disagreement after adjusting for all covariates.

**Table 6.**
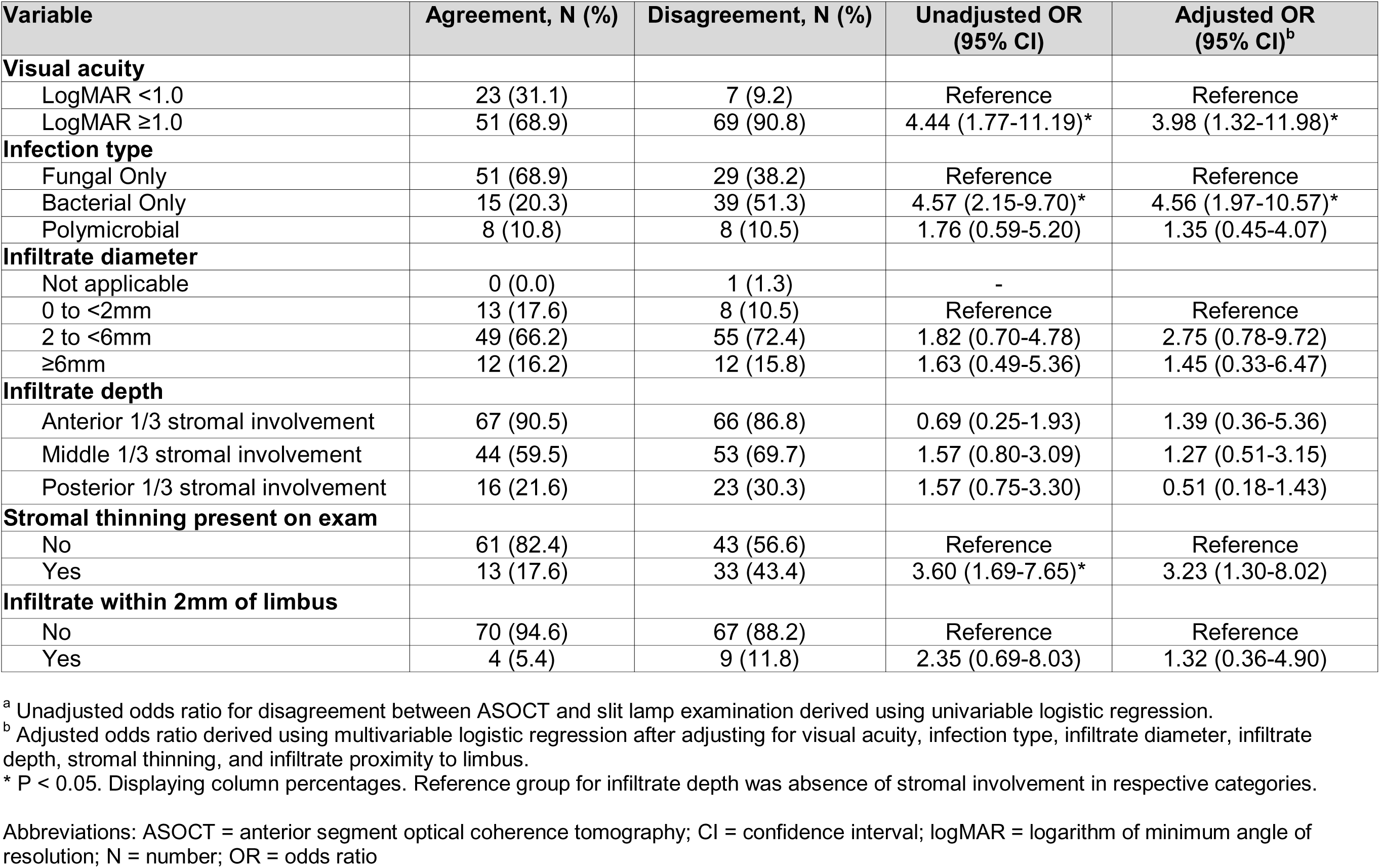
Odds of Disagreement for Any Endothelial Plaque Detection Between ASOCT and Slit Lamp Examination.

| Variable | Agreement, N (%) | Disagreement, N (%) | Unadjusted OR (95% CI) | Adjusted OR (95% CI)<sup>b</sup> |
| --- | --- | --- | --- | --- |
| **Visual acuity** |  |  |  |  |
| LogMAR <1.0 | 23 (31.1) | 7 (9.2) | Reference | Reference |
| LogMAR ≥1.0 | 51 (68.9) | 69 (90.8) | 4.44 (1.77-11.19)* | 3.98 (1.32-11.98)* |
| **Infection type** |  |  |  |  |
| Fungal Only | 51 (68.9) | 29 (38.2) | Reference | Reference |
| Bacterial Only | 15 (20.3) | 39 (51.3) | 4.57 (2.15-9.70)* | 4.56 (1.97-10.57)* |
| Polymicrobial | 8 (10.8) | 8 (10.5) | 1.76 (0.59-5.20) | 1.35 (0.45-4.07) |
| **Infiltrate diameter** |  |  |  |  |
| Not applicable | 0 (0.0) | 1 (1.3) | - |  |
| 0 to <2mm | 13 (17.6) | 8 (10.5) | Reference | Reference |
| 2 to <6mm | 49 (66.2) | 55 (72.4) | 1.82 (0.70-4.78) | 2.75 (0.78-9.72) |
| ≥6mm | 12 (16.2) | 12 (15.8) | 1.63 (0.49-5.36) | 1.45 (0.33-6.47) |
| **Infiltrate depth** |  |  |  |  |
| Anterior 1/3 stromal involvement | 67 (90.5) | 66 (86.8) | 0.69 (0.25-1.93) | 1.39 (0.36-5.36) |
| Middle 1/3 stromal involvement | 44 (59.5) | 53 (69.7) | 1.57 (0.80-3.09) | 1.27 (0.51-3.15) |
| Posterior 1/3 stromal involvement | 16 (21.6) | 23 (30.3) | 1.57 (0.75-3.30) | 0.51 (0.18-1.43) |
| **Stromal thinning present on exam** |  |  |  |  |
| No | 61 (82.4) | 43 (56.6) | Reference | Reference |
| Yes | 13 (17.6) | 33 (43.4) | 3.60 (1.69-7.65)* | 3.23 (1.30-8.02) |
| **Infiltrate within 2mm of limbus** |  |  |  |  |
| No | 70 (94.6) | 67 (88.2) | Reference | Reference |
| Yes | 4 (5.4) | 9 (11.8) | 2.35 (0.69-8.03) | 1.32 (0.36-4.90) |

<sup>a</sup> Unadjusted odds ratio for disagreement between ASOCT and slit lamp examination derived using univariable logistic regression.
<sup>b</sup> Adjusted odds ratio derived using multivariable logistic regression after adjusting for visual acuity, infection type, infiltrate diameter, infiltrate depth, stromal thinning, and infiltrate proximity to limbus.
\* P < 0.05. Displaying column percentages. Reference group for infiltrate depth was absence of stromal involvement in respective categories.
Abbreviations: ASOCT = anterior segment optical coherence tomography; CI = confidence interval; logMAR = logarithm of minimum angle of resolution; N = number; OR = odds ratio

## DISCUSSION

This study demonstrates that ASOCT provides highly reliable and sensitive detection of endothelial plaques in microbial keratitis, with near perfect inter-grader agreement for both round and flat plaque morphologies. In a rural population presenting with severe keratitis and a high predominance of fungal infections, we found that in-person ophthalmologist slit lamp examination substantially under-detects endothelial plaques, with in-person sensitivity of only 16% for round plaques, 6% for flat plaques, and 10% for either morphology compared to ASOCT. Given the strong association of endothelial plaque with infection severity and adverse outcomes,^6,7^ these findings have important implications for clinical management and outcome assessment in microbial keratitis.

During detailed ASOCT image review of 900 scans from 150 eyes with bacterial and/or fungal keratitis, our ophthalmologist graders identified two morphologically distinct subtypes of endothelial plaques that merit detailed description and separate consideration. Round endothelial plaques appeared as discrete elevated deposits on the posterior corneal surface, characterized by rounded curvature, slightly indistinct lesion edges, and deeper invasion into the anterior chamber (**Figure 1A**). In contrast, flat plaques presented as linear hyperreflective deposits along Descemet membrane with sharper edges, a thinner profile with minimal anterior chamber protrusion, and a broader plane of attachment along the corneal endothelium (**Figure 1B**). Both lesion types can occur in the same eye (**Figure 1C and 1D**), and are often accompanied by hyperreflectivity of Descemet membrane well beyond the area of infection. These distinct morphological patterns may represent different stages of disease progression or fundamentally different pathophysiological processes and warrant further exploration in future clinical and histopathologic studies. The clinical significance of distinguishing these morphologies is supported by our finding that they exhibit different detection characteristics. Round plaques showed higher clinical detection sensitivity (16.3%) compared to flat plaques (6.2%), suggesting that the three-dimensional prominence of round plaques and their invasion into the anterior chamber makes them more visible on slit lamp examination compared to the subtler layering of flat plaques which can be more easily overlooked. Both morphologies, however, were dramatically under-detected by clinical examination compared to ASOCT. Given the association between endothelial plaques and anterior chamber organism invasion in fungal keratitis,^6,8^ the morphological distinction between round plaques with deeper invasion and flat plaques with broader surface attachment may have implications for treatment decisions and prognosis.

**Figure 1.**
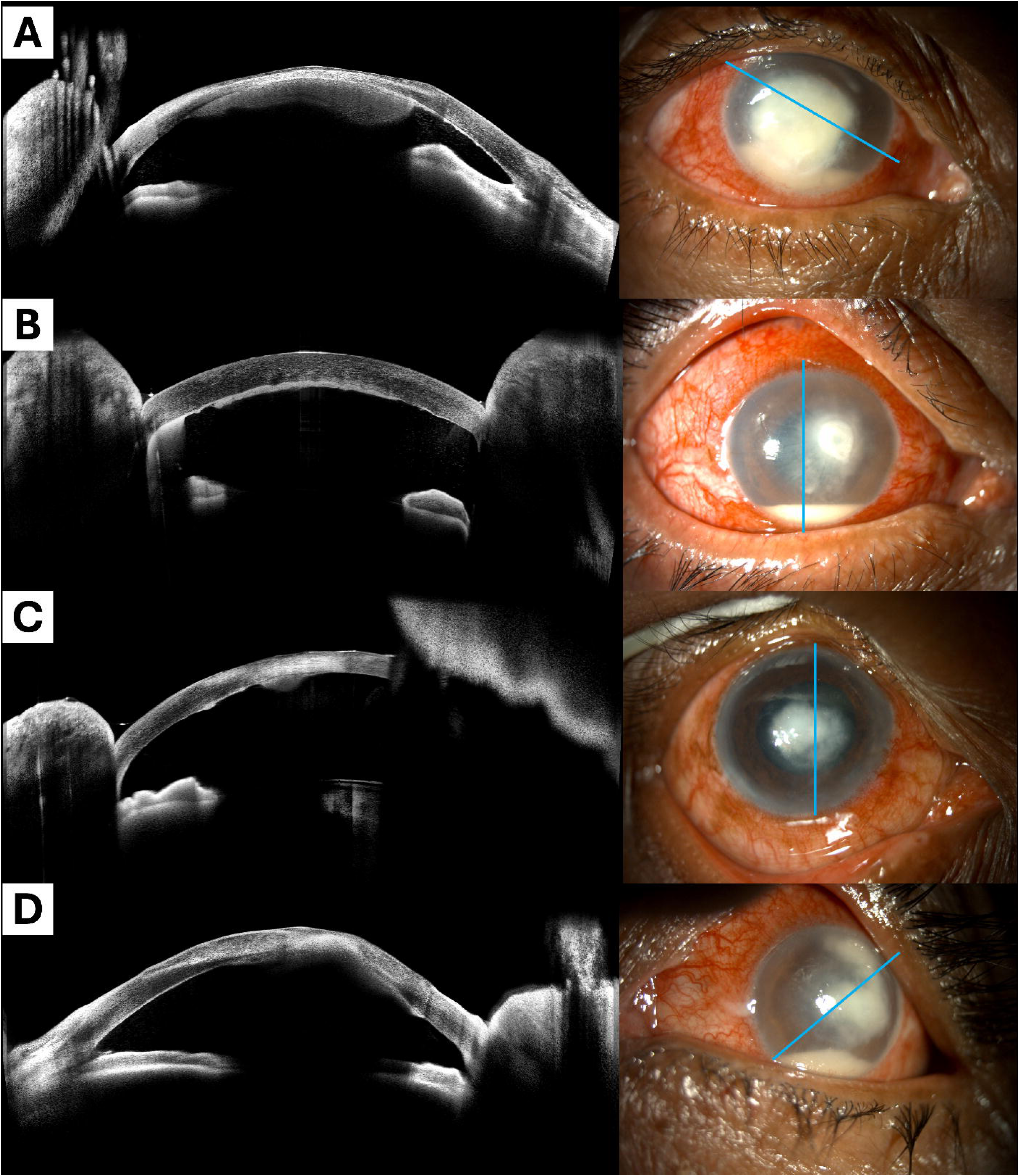
Representative ASOCT images demonstrating distinct endothelial plaque morphologies. **(A)** Round endothelial plaque: Elevated hyperreflective deposit on the posterior corneal surface with rounded curvature, indistinct lesion edges, and deeper invasion into the anterior chamber. **(B)** Flat endothelial plaque: Hyperreflective sheet-like linear deposit along Descemet membrane with sharper margins, thinner profile with minimal anterior chamber protrusion, and broader plane of attachment to the endothelium. The flat endothelial plaque is distinct from the hypopyon visible in the inferior angle. **(C)** Combined lesion showing features of both round and flat endothelial plaque morphologies in the same eye. **(D)** Combined lesion with round endothelial plaque underneath corneal apex, as well as linear deposits and hyperreflectivity along the Descemet membrane.

The near perfect inter-grader agreement for ASOCT detection across all morphologies (kappa 0.84-0.88) supports use of this imaging modality as a readily standardizable assessment tool in microbial keratitis research. Slit lamp grading of endothelial plaques is inherently subjective and depends on adequate visualization through an often opaque, edematous cornea. ASOCT provides objective cross-sectional imaging that is less affected by anterior corneal opacity, enabling more consistent assessment across examiners and study sites. Notably, the composite outcome (either morphology) showed the highest intra-grader reproducibility for both graders, suggesting that assessing for presence of any endothelial plaque may be the most reliable approach for clinical evaluation and research.

The substantial discrepancy between ASOCT and clinical detection (55% versus 6% graded as having any endothelial plaque) highlights a significant limitation of traditional slit lamp assessments of microbial keratitis. Clinical examination detected less than one-sixth of ASOCT-identified round plaques and less than one-tenth of flat plaques. Under-detection was most pronounced in eyes with poor visual acuity (which correlated with more severe disease) and for detection of either type of endothelial plaque, in bacterial infections. The high specificity of clinical examination (>94-99% depending on morphology) suggests that when clinicians do identify endothelial plaques on slit lamp biomicroscopy, these lesions are truly present and rather large in size. However, absence of visible endothelial plaque on slit lamp examination cannot reliably exclude its presence.

Compared to eyes with fungal keratitis, eyes with bacterial keratitis had 4.6-fold higher odds of disagreement in detection of endothelial plaque on ASOCT versus slit lamp examination. Interestingly, this association was strongest for the composite outcome of “any endothelial plaque” rather than individual round or flat plaque morphologies, suggesting that bacterial keratitis may be associated with under-detection of both plaque subtypes. Potential reasons for increased endothelial plaque detection on ASOCT in bacterial versus fungal keratitis could include increased stromal infiltrate size, increased infiltrate density, or other changes in the inflammatory pattern for bacterial infections that could affect endothelial plaque formation and visibility. Microbiology scraping was performed at the time of initial clinical assessment and immediately submitted for Gram stain and potassium hydroxide prep. Smear results usually became available within an hour, were shared with the ophthalmologist at the time of final patient assessment, and could therefore have influenced clinicians’ slit lamp assessments. Since endothelial plaques are more commonly associated with fungal infections, clinicians evaluating eyes with microbiologically confirmed bacterial keratitis could have had a lower index of suspicion for identifying endothelial plaque among bacterial cases compared to confirmed fungal infections. The observed association warrants further investigation into possible causes as well as replication in other patient populations.

Our results may support incorporation of ASOCT into standardized imaging protocols for assessing disease severity and treatment response in microbial keratitis, particularly if future research demonstrates an association between endothelial plaque size or morphology and clinical outcomes. In particular, identification of endothelial plaques, characterization of their morphology, and measurement of their size may inform treatment approaches including timing of surgical intervention. In geographically isolated or resource-limited settings where cornea specialists may not be available for in-person examination, telemedicine models using ASOCT paired with remote ophthalmologist assessment could expand access to expert assessment closer to patients’ homes.

### Strengths and Limitations

Strengths of this study include the large sample size, microbiological confirmation of all bacterial or fungal keratitis cases, standardized imaging and data collection protocol, masked independent grading by ophthalmologists, formal assessment of both intra- and inter-grader reliability, and systematic evaluation of two distinct plaque morphologies. The study population reflects a real-world clinical cohort from an endemic region with substantial representation of both fungal and bacterial keratitis.

Several limitations warrant consideration. This single-center study from rural India may not generalize to all clinical settings or patient populations, particularly those with different distributions of causative organisms such as viral or parasitic keratitis. The patient population at SNC Chitrakoot presented with more severe cases of microbial keratitis than many other centers, with nearly 60% of eyes presenting with visual acuity of counting fingers or worse and a median of 15 days elapsing between symptom onset and initial patient presentation to the tertiary care center. Although we used remote ASOCT grading as the reference standard rather than histopathological confirmation, some ASOCT findings classified as endothelial plaque may represent other pathology, and the morphological distinction between round and flat plaques has not been validated histologically or via longitudinal outcomes studies. In-person slit lamp examination recorded only “endothelial plaque” without distinguishing the round and flat morphological subtypes of endothelial plaque identified on ASOCT, limiting head-to-head analyses of clinical detection by morphologic subtype. ASOCT imaging with the Anterion Metrics app captures six cross-sections of each eye, but these scans may miss lesions outside the scan plane. Future studies should evaluate whether multiple scan locations improve detection sensitivity even further.

## CONCLUSION

In conclusion, ASOCT enables highly reliable and sensitive detection of endothelial plaques in microbial keratitis, with substantial to near perfect intra- and inter-grader agreement for both round and flat endothelial plaque morphologies. In-person slit lamp examination substantially under-detects endothelial plaques compared to ASOCT imaging. These findings support incorporation of ASOCT into clinical assessment and research protocols for microbial keratitis.

## Data Availability

All data produced in the present study may be available upon reasonable request to the authors and in accordance with local regulations.

## ACKNOWLEDGMENTS

The authors thank the clinical and research staff at SNC Chitrakoot for their assistance with patient recruitment and data collection.

